# Pre-treatment EEG microstates and response to antipsychotic drugs in first episode psychosis

**DOI:** 10.64898/2026.09.26.26364042

**Authors:** Jose M Rubio, Michael Murphy, Wadim Vodovozov, Ricardo Carrion, Todd Lencz, Mate Baradits, Dost Ongur, Mehrdad Sadri, John M Kane, Anil K Malhotra

## Abstract

**Background and Hypothesis:** First-line antipsychotics are ineffective in ∼40% of people with first episode psychosis (FEP). Electroencephalogram (EEG)-based predictive biomarkers could be a scalable solution to reduce trial-and-error.

**Study Design:** We acquired EEG in 56 minimally treated FEP individuals before treatment initiation, and 34 matched healthy controls (HC). Treatment response was defined as >50% improvement in BPRS scores by 12 weeks of treatment. Modified k-means clustering of resting-state EEG yielded six microstates including canonical topographies (A-D). The first principal component (PC1) of microstate properties derived from HCs served as a normative reference onto which FEP patients were projected. Baseline PC1 was used to predict treatment response in adjusted regression analyses. To test stability of results, a second dataset of HCs (n=92) was fitted using the same microstate solution and compared to our native HC dataset. PC1 scores derived from this independent normative dataset were also used for predicting response in FEP.

**Study Results:** Individual microstates replicated established patient-control differences (e.g., higher presence of microstate C compared to D, p<0.001). PC1 explained 34.8% of variance. Baseline PC1 predicted subsequent treatment response (adjusted OR=0.63, p<0.001), yielding AUC=72.3% in leave-one-out cross-validation (p_perm_<0.001) when combined with clinical information. PC1 from our HC transported well to the second healthy control EEG dataset (n=92) (r=0.9,p<0.001; r=0.89,p<0.001 ). PC1 in FEP derived using PC1 weights from a second normative dataset also predicted response in FEP (OR=0.70, p=0.03).

**Conclusions:** EEG microstate architecture differing from the expected norm is associated with lower odds of treatment response in FEP.

## Introduction

First-line antipsychotic drugs are ineffective for approximately 40% of individuals with a first episode of psychosis (FEP)[1]. This often leads to a trial-and-error process in the search for effective medication. A particularly concerning scenario is when trial and error leads to long delays in initiating clozapine, the only drug approved for treatment resistance [2]. Median delays are approximately 5 years [3], and the later treatment is introduced, the less likely it is to be effective[4]. Therefore, there is a great need to transition from trial-and-error treatment selection to personalized medicine to optimize patient experience and treatment outcomes.

In a recent review, we examined biomarker readiness for use in clinical practice[5]. fMRI predictive biomarkers were considered relatively advanced[6]. One obstacle they face though is scalability, since they may not be accessible to most patients experiencing FEP. This has called for exploration of alternatives to fMRI for predictive biomarker development.

Electroencephalography (EEG) has several scalability advantages[7]. EEG measures transient, quasi-stable patterns of scalp electrical activity that reflect the dynamic spatial organization of brain activity. It has a favorable balance of temporal resolution, cost-effectiveness [8]. EEG has been successfully deployed in low-resource rural communities for some neurological applications[9].

EEG microstates are defined as quasi-stable topographical patterns of electrical activity lasting approximately 60-120 milliseconds before rapidly transitioning to a different configuration[8]. The canonical microstates (typically labeled A through D, with some studies also identifying E and F) represent specific topographical configurations that are highly replicable across studies[10, 11]. These canonical microstates are thought to reflect the momentary synchronization of large-scale brain networks[10, 11].

In schizophrenia, meta-analyses have identified consistent alterations, most robustly increased microstate class C and decreased microstate class D, specifically, class C is more present/occurs more frequently and class D is shortened/less present in schizophrenia patients versus controls[12–14]. More broadly, microstate characteristics have been associated with transition from the prodrome to full-blown psychosis [15–18], psychosis severity including auditory verbal hallucinations [19], and global functioning [20]. Given their abnormalities in unaffected siblings, microstates have been proposed as candidate endophenotypes [14].

Two studies have examined the association between microstates and treatment response. Kikuchi et al. [21], in a small sample (n=14) of multi-episode patients with schizophrenia, found that changes in overall microstate duration over 4 weeks of treatment strongly correlated with symptom improvement (r=−0.689, p=0.005). Specifically, change in the mean duration of microstate D was most associated with symptom reduction (r=−0.709, p=0.003). More recently, De Pieri et al. [22] examined EEG microstates in 40 multiepisode patients with acute-phase schizophrenia in relation to their treatment response 6 weeks after baseline. This work revealed significant baseline differences in microstate C, D and E between individuals who later responded to antipsychotic and those who would not. These preliminary data suggest that EEG microstate capture biological processes that are relevant for antipsychotic treatment response, opening a new avenue for biomarker development. The next step in this line of research is to replicate these findings in minimally treated FEP – the application for which a predictive biomarker would be most useful – under standardized treatment conditions, as well as to test whether differences in overall microstate architecture, rather than isolated specific properties, carry prognostic variance. .

Here, we aim to examine microstate properties as markers of treatment response in first-episode psychosis using a normative modeling framework. We first establish normative patterns of microstate organization in healthy controls using PCA, then project patients onto this normative space to derive a single, interpretable composite measure (PC1). Based on previous literature implicating various microstate features in treatment response, we hypothesize that baseline deviation from normative microstate organization (PC1) will predict response to 12 weeks of standardized antipsychotic treatment in first-episode psychosis.

## Patients and Methods

To test our hypothesis, we collected data from a clinical trial of antipsychotics (risperidone or aripiprazole) in FEP. EEG was acquired pre-treatment and after 12 weeks of treatment, and psychopathology ratings were acquired multiple times through the course of the trial. EEG was also acquired in 34 healthy controls.

### Research Participants And Clinical Trial

We recruited participants who presented to their first treatment for psychosis at the Zucker Hillside Hospital with <2 weeks of cumulative lifetime exposure antipsychotics (43% treatment naïve, median exposure = 5 days). All participants were between 18 and 40 at the time of enrollment. Inclusion criteria were DSM-5 defined diagnosis of schizophrenia, schizophreniform, schizoaffective disorder, psychosis not otherwise specified or Bipolar I (acute manic or mixed episode). Diagnostic eligibility was confirmed by the Structured Clinical Interview for DSM-5 (SCID-5)[23]. Exclusion criteria included for all participants included: 1. Any medical condition or treatment known to affect the brain; 2. Any medical condition which requires treatment with a medication with psychotropic effects; 3. Cognitive or language limitations that would preclude subjects providing informed consent; 4. Active substance dependence (within the last 6 months) exclusive of cannabis or nicotine; 5. Loss of consciousness for longer than 10 min or with neurological sequela; and 6. History of Electro Convulsive Treatment. Healthy controls met the same criteria other than not meeting DSM-5 criteria at the time of the assessment of any psychotic, affective, anxiety or drug use disorder. Patients underwent a standardized flexible-dose treatment protocol with risperidone or aripiprazole for 12 weeks, and regular clinical ratings. Patients underwent assessment by the Brief Psychopathology Rating Scale (BPRS-A)[24] at baseline and weeks 1, 2, 3, 4, 6, 8, 10, 12. Consistent with the FEP literature, we defined treatment response as a reduction of symptoms >50% between baseline and predicted week 12 scores [25]. All participants provided written informed consent under a protocol approved by the Institutional Review Board (IRB) of the Feinstein Institutes for Medical Research at Northwell Health, and the data were part of NIH funded project R01MH108654.

### EEG Acquisition

EEG was acquired for each participant prior to starting the antipsychotic trial and once again after completing 12 weeks of treatment. For each participant and timepoint, 10 min of eyes-closed resting EEG data was collected using an EEG cap with 64 A g–AgCl electrodes sampled at 1024 Hz with a common vertex reference. EEG data was bandpass filtered from 0.5 to 40 Hz. Bad channels were identified by an automated threshold-based procedure part of the CARTOOL software package which calculates several statistics on each channel at each time point and then labels as bad channels that are: a) flat, b) excessively noisy, or c) out of scale with the other channels[26], and confirmed with visual inspection and then interpolated with spherical splines. Segments of data with EEG artifact were excluded from further analyses. ICA removal of eye blinks was not conducted as eyes closed data are largely robust to ICA eye blink removal provided artifactual segments of data are removed[27]. Data was average referenced and downsampled to 128 Hz. Data analyses were performed in MATLAB (MathWorks, Natick MA) and CARTOOL [26]. Minute-by-minute analysis of alpha activity using a bandpass filter indicated that patients remained awake during the recording period (i.e., <50% of epoch occupied by alpha)(, no recordings were discarded due to drowsiness. For each channel, the power spectra distribution was calculated using Welch’s method with Hamming windows of 2 s2□s in length, 50% overlap, and then averaged across channels [28] (Supplementary Figure 1). Acquisition of FEP patients and matched healthy controls was run in parallel as part of project R01MH108654 with identical approach.

### Microstate Analyses

Microstate analysis was performed using CARTOOL, a powerful standalone software suite for advanced analysis and visualization of multichannel EEG [26]. EEG data was spatially filtered by replacing the data from each electrode with a weighted inter-septile mean of that electrode and its six nearest neighbors [29].

To generate microstate topographies, we used a two-step segmentation approach. We limited our segmentation analysis to time points corresponding to local maxima of the global field power[30]. For each participant, we randomly selected 20 subsets of the data that together covered at least 95% of the original set. For each subset, we applied a polarity-insensitive k-means analysis with k ranging from 1 to 10 [31]. The optimal number of microstates was selected using the meta-criterion implemented in CARTOOL, which combines seven independent optimization criteria: Krzanowski-Lai index, Cross-Validation, Davies-Bouldin index, Point-Biserial, Dunn index, Silhouette index, and Gamma index [32]. The optimal k is selected as the value that maximizes the median rank across these criteria. The resulting topographies were saved for each subset. Next, we ran another polarity-insensitive k-means analysis with k = 1 to 10, this time using the outputs from the first round of segmentation as input. To reduce type I error rates, we used the same combined set of topographies for patients and controls [33]. This combined set of microstate topographies was fitted back to the original EEG data. Microstate segments less than 15 ms in duration were rejected[34]. This analysis produced a microstate label for each time point in the original EEG data. Time points that were poorly correlated with any microstate topography (defined as |correlation| *<* 0.5) were not included in the analyses.

Microstate coverage was defined as what portion of the EEG data was assigned to each microstate, duration was defined as the average length of time a given microstate persisted in ms, explained variance was defined as how much of the variance in the EEG data was accounted for by each microstate, and occurrence which refers to the proportion of time that a specific microstate configuration occurs within a given EEG recording.

Microstate templates were derived from the patient sample using unsupervised k-means clustering on topographical patterns, without reference to treatment outcome labels, and subsequently applied to healthy controls. This approach ensures that the spatial configurations captured are relevant to the clinical population of interest, consistent with recommendations to derive templates from the sample in which microstate alterations are hypothesized[33].

While this means the feature space is defined by patient topographies, the temporal statistics extracted under these templates are computed independently for each subject from their own EEG data. The templates define which spatial configurations to quantify — analogous to an anatomical atlas defining regions of interest — but do not determine the temporal statistics that serve as predictors. To further rule out pipeline-specific artifacts, we conducted a transportability analysis using an independent healthy control dataset (see below).

A 6 microstates solution was favored by the meta-criterion selection procedure, which included the canonical microstates A, B, C, D, as well as previously described frontocentral distribution microstate E and posterior-central microstate F (Figure 1), following the labels of the most recent review [11].

**Figure 1.**
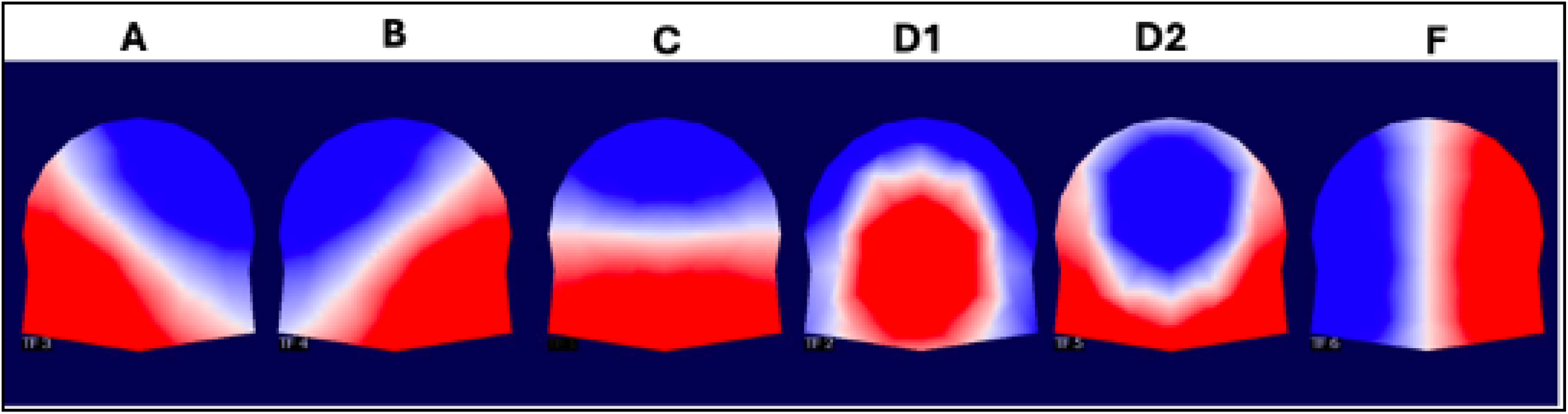
Six microstate solution found in cohort of first episode patients before treatment onset Legend: Scalp topographies of the six microstates identified across FEP. Topographies correspond with Microstates: A (right-posterior to left-anterior diagonal), B (left-posterior to right-anterior diagonal), and C (frontal-to-occipital anterior-posterior orientation), D1 and D2 (frontocentral), and F (right-lateralized posterior-to-anterior distribution). Red and blue indicate opposing polarities.

### Dimensionality Reduction: Principal Component Analysis On Healthy Controls And Projection Of Patients Onto Normative Space

By using Principal Components Analysis (PCA) from healthy controls and projecting the weights onto FEP participants microstate properties, we solve two problems. One is the multicollinearity, almost by definition, between microstate properties (Supplementary Figure 2). Each principal component (PC) often captures more variance than a single raw microstate property[35], allowing models with fewer variables which have less risk of overfitting. The second is that meaningful predictive features may not largely overlap between patients[36]. For example, in one patient the prediction may be driven by short duration of microstate D, while in other it could be excessive occurrence. By using deviations of the normative architecture as primary metric, the question becomes not what specific microstate property predicts prognosis, but rather whether a deviation from a normative architecture (which could be in different aspects between patients), is predictive.

PCA was performed exclusively on matched healthy control data (n = 34) to establish a normative reference space. We first standardized (z-scored) the 24 microstate features that would go into the model. In the decomposition, the first principal component (PC1) explained 34.8% of the variance (Eigenvalue=8.61), and >80% of the variance was explained by the first four principal components (87.8%; Supplementary Figure 3). We then evaluated the stability of the resulting components using bootstrap resampling (2,000 iterations). For each bootstrap sample, we recomputed the PCA and calculated the cosine similarity between bootstrap loadings and the original loadings, with sign correction applied to account for arbitrary sign flips. Component stability was defined as ≥70% of bootstrap resamples achieving cosine similarity > 0.80 with the reference loadings. For PC1 the median cosine over bootstraps was=0.95 with 70.2% of the bootstraps being >0.8, demonstrating stability, while the rest of the PCs did not meet our stability criteria, except for the fifth PC (Supplementary Figure 4). The loadings onto PC1 were dominated by microstate C and F being A the least influential (Supplementary Figure 5). As additional analyses, we explored the distributions of the projected PC2 to PC5 onto patients, to test whether they carry a meaningful prognostic signal. Only PC2 showed a significantly different distribution between responders and non-responders (Supplementary Figure 6). Furthermore, to assess whether our results were contingent on the specific healthy control sample used to derive PC1, we conducted a transportability analysis. An independent dataset of healthy controls from the LEMON study (n=92)[37] was used as a second normative reference. This dataset was collected by the Max Planck Institute and consists of n=153 (92 with valid EEG data) HC participants with 25.1±3.1 years, range 20–35 years, 45 female, collected between 2013 and 2015 to study mind-body-emotion interactions. EEG from the LEMON participants were fitted using the same six-microstate solution. PCA was performed independently on this new sample, and also cross-projected between both HC datasets. Resulting PC1 scores were compared using score agreements between lemon own PC1 scores and our HC PC1 weights projected onto the LEMON microstate properties (PC1 r=0.9, PC2 r=0.86, PC3 r=0.49, PC4 r=0.84, PC5 r=0.96) (Supplementary Figure 7). Given the signs of instability of PC2, and the relatively minimal variance explained by PC5 (5.1%), we favored retaining only PC1 for the primary analyses as best compromise between simplicity and explanatory power. PC2 to PC5 were kept for exploratory analyses.

Patient microstate properties were standardized, then projected onto the PC1 axis defined by our HCs. This projection yields a PC1 score for each patient representing their deviation from normative microstate organization and were used for the primary analyses.

### Statistical analyses

We first examined the 24 baseline microstate parameters across groups (healthy controls, responders, non-responders) using boxplots and calculated pairwise effect sizes (Cohen’s d) and FDR corrected p-values.

To test whether baseline deviation from normative microstate organization predicts treatment response, we conducted logistic regression with treatment response (responder vs. non-responder) as the dependent variable and baseline PC1 as the primary predictor. Covariates included age, sex, baseline BPRS total score, medication type (risperidone vs. aripiprazole), diagnostic category (non-affective vs. affective psychosis), and treatment-naïve status. We report odds ratios with 95% confidence intervals derived using the profile likelihood method. To evaluate predictive performance, we conducted leave-one-out cross-validation (LOO-CV). For each fold, the held-out subject’s data was excluded, the remaining data were standardized, and a logistic regression model was fit to predict response. The predicted probability for the held-out subject was recorded. This procedure was repeated for all subjects. We computed the area under the receiver operating characteristic curve (AUC) from the LOO-predicted probabilities, along with accuracy, sensitivity, and specificity at the optimal threshold. To assess whether predictive performance exceeded chance, we conducted permutation testing (1,000 permutations) in which response labels were randomly shuffled and the entire LOO-CV procedure was repeated, generating a null distribution of AUCs. The permutation p-value was calculated as the proportion of null AUCs exceeding the observed AUC. We conducted two LOO-CV models: 1) PC1 alone, and 2) PC1 plus covariates (age, sex, baseline BPRS, medication, diagnosis, treatment-naïve status), to evaluate the incremental value of clinical covariates over the EEG-based predictor. In addition, to measure the consistency of our results, we measured treatment response as continuous measure of symptom change over the 12 weeks of treatment. For this, we ran a linear regression adjusting for age, sex, baseline BPRS, and treatment-naïve status of baseline PC1 over BPRS percent change at week 12.

Finally, we tested whether PC1 individual scores in FEP derived from projecting the LEMON PC1 weights, were used in the same models as described above to test whether they also were predictive of treatment response.

The Python scripts used for these analyses are publicly available at https://github.com/lorente01/microstates The study was registered as a clinical trial at https://clinicaltrials.gov/study/NCT02822092

## Results

### Clinical characteristics

Out of 56 participants for whom EEG were collected at baseline n=32 (57.14%) met response criteria by week 12. At baseline, mean age was 23.5 (SD=5.28) years old, n=35 (62.5%) were male. Fifteen (26.7%) participants were white, 24 (42.8%) were black, and the remainder of the cohort were other race. Baseline mean BPRS Total score was 45.12 (SD=6.39) and mean CGI 4.89 (SD=0.56). Only n=6 (10.7%) were diagnosed with bipolar I disorder with psychotic features, and the rest n=50 (89.3%) with non-affective psychoses. Forty participants (71.42%) were treated with risperidone, while the rest were treated with aripiprazole. There were no statistically significant differences between FEP and HC (Table 1).

**Table 1.** Clinical characteristics.

|  | All Patients<br>(n=56) | Non-Responders<br>(n=24) | Responders<br>(n=32) | p-value (R<br>vs NR) | Healthy Controls<br>(n=34) | p-value<br>(Patients<br>vs HC) |
| --- | --- | --- | --- | --- | --- | --- |
| Age | 23.49 (5.42) | 24.28 (6.80) | 22.90 (4.12) | 0.38 | 25.32 (4.8) | 0.09 |
| Male | 35 (62.5%) | 14 (58.3%) | 21 (65.6%) | 0.78 | 18 (52.9%) | 0.37 |
| Black race | 26 (46.4%) | 11 (45.8%) | 15 (46.9%) | 0.6 | 12 (35.9%) | 0.29 |
| Hispanic ethnicity | 15 (26.8%) | 8 (33.3%) | 7 (21.9%) | 0.6 | 10 (29.4%) | 0.79 |
| Baseline BPRS | 44.84 (6.39) | 44.71 (5.44) | 44.94 (6.53) | 0.51 | N/A | N/A |
| Non affective psychosis | 50 (89.3%) | 22 (91.7%) | 28 (87.5%) | 0.95 | N/A | N/A |
| Risperidone | 40 (71.42%) | 17 (70.8%) | 23 (71.9%) | 0.93 | N/A | N/A |
| Aripiprazole | 16 (28.57%) | 7 (29.2%) | 9 (28.1%) | 0.93 | N/A | N/A |

### Distribution of microstate features by group

At the group level, 22 out of 24 microstate properties were significantly different between HC and non-responders after correcting for multiple comparisons. For the comparison between HC and responders it was 18, and between responders and non-responders it was 3. Group level distribution of microstate features are described in detail in Figure 2 and Supplementary Figure 8.

**Figure 2.**
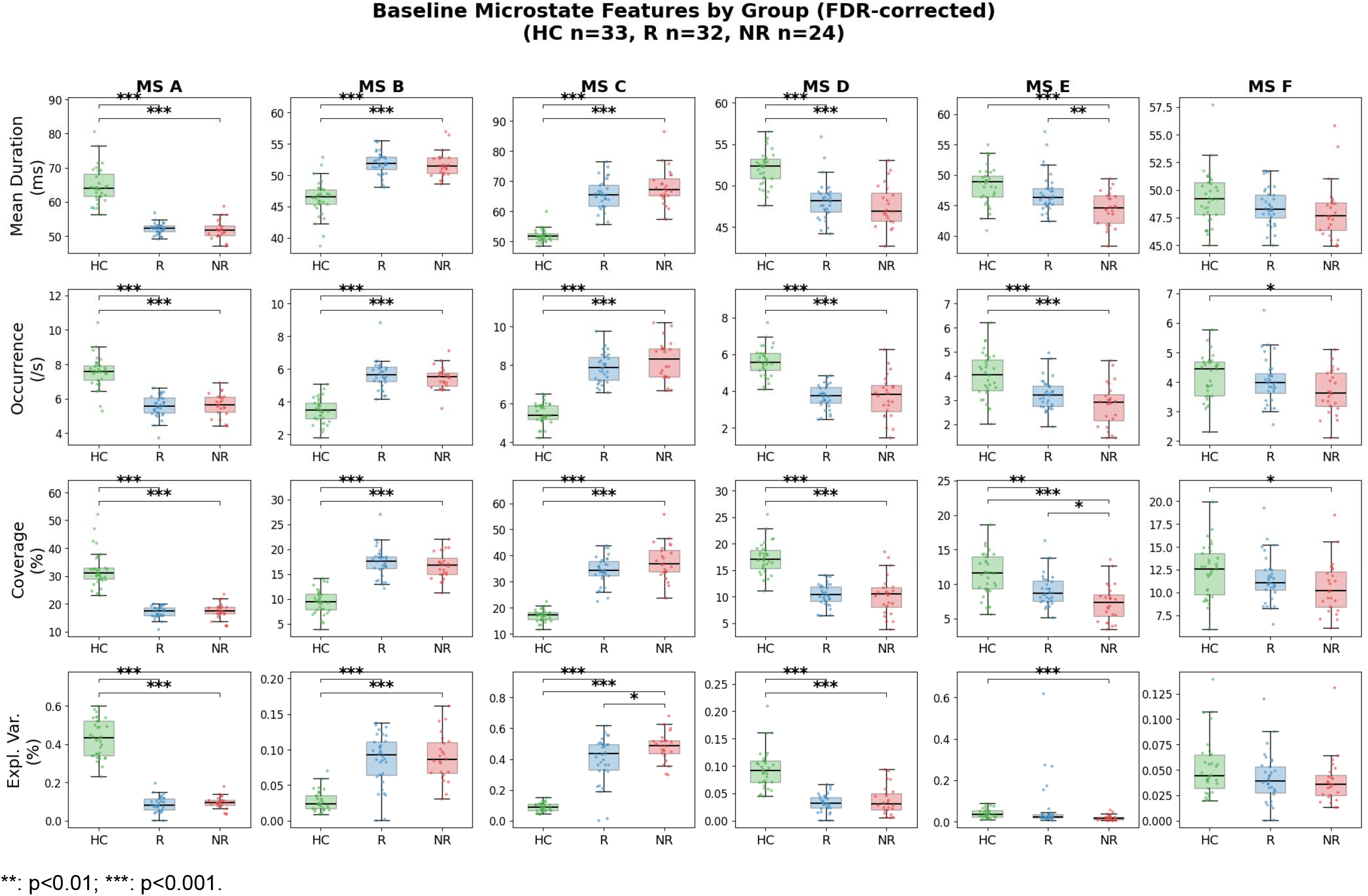
Distribution of microstate features by group at baselin.

### Transportability of PC1 as normative framework

The correlation between native PC1 scores in the LEMON participants and the PC1 scores in LEMON participants using the ZHH HC PC1 weights was r=0.9, p<0.001. In the other direction, ZHH HC participants had PC1 native individual scores that correlated at r=0.89, p<0.001 with PC1 individual scores using the LEMON weights (Figure 3).

**Figure 3.**
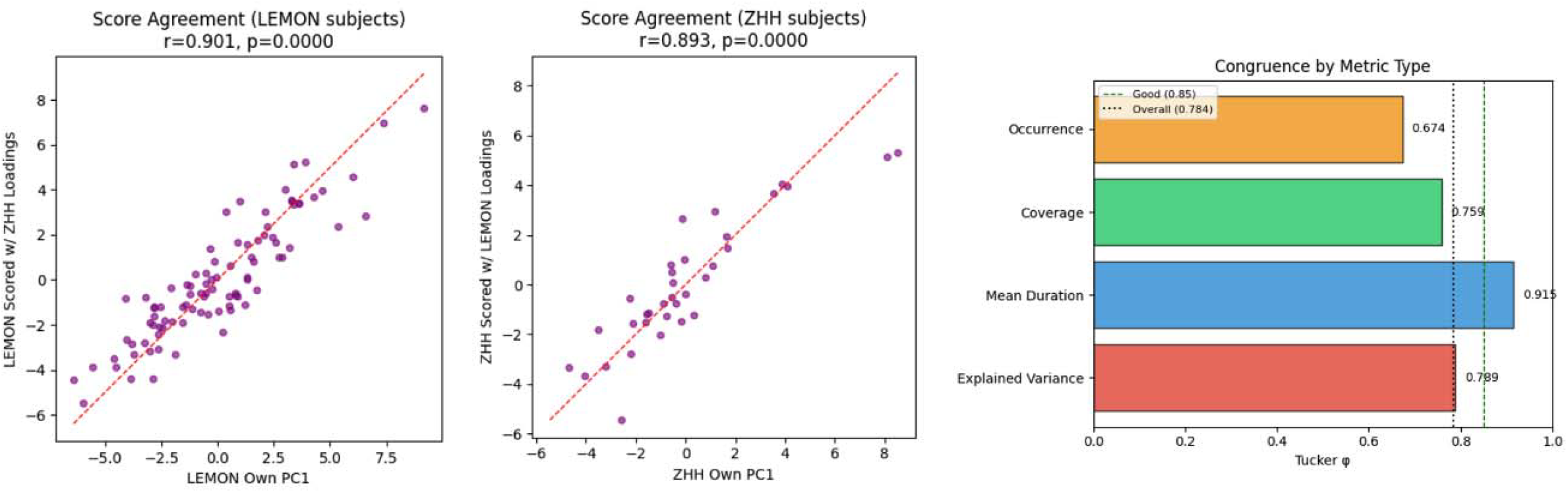
Bidirectional cross projection of PC1 scores between our native healthy control sample and the publicly available.

### Baseline PC1 and treatment response

At the PC1 level, non-responders showed significantly higher PC1 scores than responders (mean difference=1.97, Cohen’s d=0.76, t=2.83, p=0.007), indicating greater deviation from normative microstate organization in patients who subsequently failed to respond to treatment. Responders did not differ from healthy controls (d=0.05, p=0.838), while non-responders showed elevated PC1 relative to healthy controls (d=0.66, p=0.017) (Figure 4A). In the multivariable logistic regression model adjusted for age, sex, baseline BPRS total score, medication type, diagnostic category (non-affective vs. affective psychosis), and treatment-naïve status, baseline PC1 significantly predicted non-response (OR=0.63 per unit increase in PC1; 95% CI=0.46–0.88; p=0.007). Baseline BPRS total score was also a significant predictor (OR=1.17; 95% CI=1.03–1.32; p=0.013), while treatment-naïve status was not (OR=0.52; 95% CI=0.12–2.18; p=0.37). The model pseudo-R^2^ was 0.276. Leave-one-out cross-validation (LOO-CV) using PC1 alone yielded AUC=0.648 (permutation p=0.013, 1,000 permutations), with accuracy=62.5%, sensitivity=78.1%, and specificity=41.7% at the optimal threshold. Adding clinical covariates (age, sex, baseline BPRS, medication, diagnosis, treatment-naïve status) improved performance to AUC=0.723 (permutation p=0.002), accuracy=75.0%, sensitivity=81.2%, and specificity=66.7%, representing a modest improvement of ΔAUC=+0.074 (Figure 4B). The significant permutation p-values indicate that both models performed above chance levels. Baseline PC1 was significantly correlated with BPRS percent change, such that higher PC1 scores (i.e., more normative microstate organization) were associated with greater symptom improvement (Spearman ρ=+0.38, p=0.004; Pearson r=+0.30, p=0.023). In linear regression adjusted for age, sex, baseline BPRS, and treatment-naïve status, baseline PC1 remained a significant predictor of symptom improvement (β=+1.49, SE=0.64, t=2.32, p=0.024), indicating that each unit increase in baseline PC1 was associated with 1.49 percentage points greater symptom reduction. Baseline BPRS was also significant (β= − 0.72, p=0.012), such that higher baseline severity predicted greater improvement, as expected. The adjusted model R^2^=0.238 (Figure 4C).

**Figure 4.**
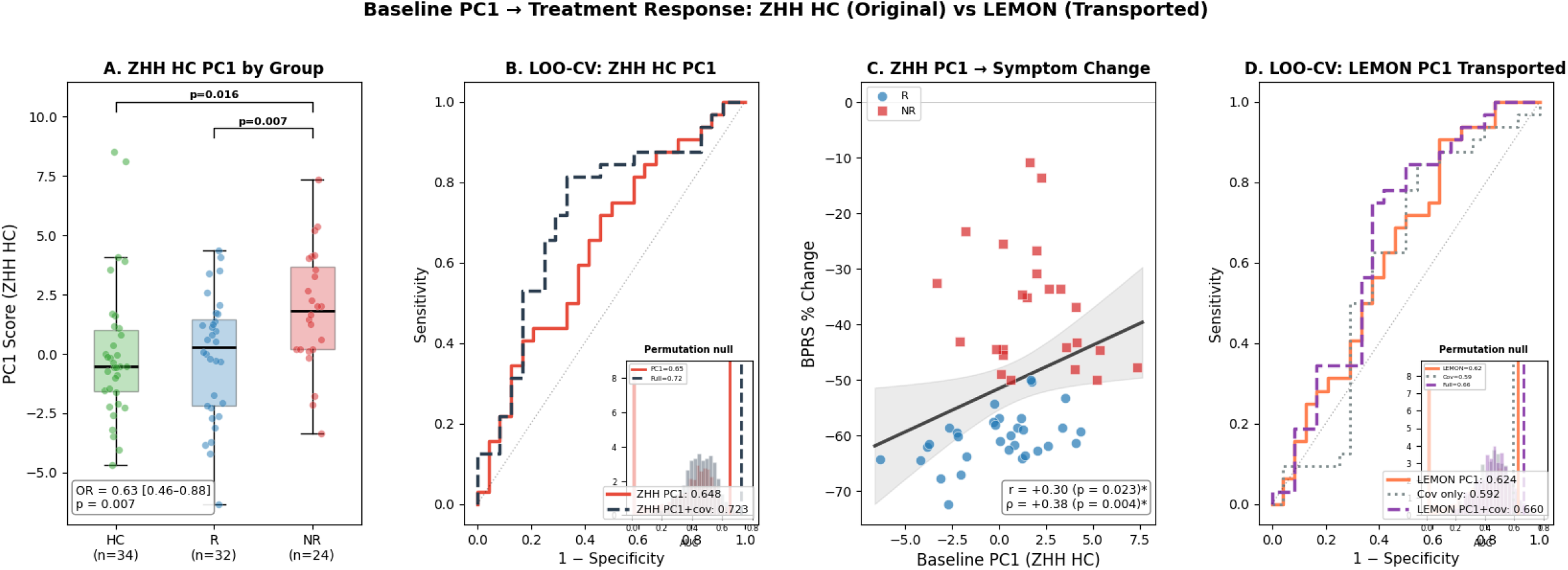
Association between baseline PC1 and antipsychotic response in first episode psychosis. A. First principal Component (PC1) scores differed across groups. Non-responders showed higher PC1 than responders (Cohen’s d = 0.76, p = 0.007), while responders did not differ from healthy controls (d = 0.05, p = 0.838). In a logistic regression adjusted for sex, age, treatment, naïve status, diagnosis, PC1 predicted response (Odds Ratio = 0.63, 95% Confidence Interval: 0.46–0.88, p = 0.007). B. Leave-one-out cross-validation using PC1 alone yielded Area Under the Curve (AUC) = 0.648 (permutation p = 0.013; accuracy = 62%, sensitivity = 78%, specificity = 42%). Adding covariates (age, sex, baseline BPRS, medication, diagnosis) yielded AUC = 0.723 (1000 permutations p = 0.002; accuracy = 75%, sensitivity = 81%, specificity = 67%) C. Baseline PC1 predicted symptom improvement dimensionally (ρ = +0.38, p = 0.004; in linear regression adjusted for sex, age, treatment, naïve status, diagnosis β = +1.49, p = 0.024). D. Leave-one-out cross-validation using PC1 from the LEMON dataset projected onto FEP, yielding Area Under the Curve (AUC) = 0.624; which when adding covariates (age, sex, baseline BPRS, medication, diagnosis) increases to 0.660

### Replication of Results Using a Normative Space Derived From a Different Healthy Control Dataset

When projecting the PC1 from the LEMON dataset (n=92) onto the FEP cohort, we obtained similar results as when using our own HC dataset. Namely, in the multivariable logistic regression OR=0.70 (95% CI=0.51-0.96; p=0.027). In the LOOCV using only LEMON Projected PC1 AUC=62.4%, while combined with clinical characteristics AUC=66%, p_perm_<0.001 (Figure 4D).

## Discussion

This study demonstrates that baseline deviation from normative microstate organization, captured by a single principal component (PC1) derived from healthy controls, predicts treatment response to first-line antipsychotics in first-episode psychosis. Non-responders showed greater deviation from normative patterns than responders, who were comparable with healthy controls. Results notably replicated when an independent normative dataset was used as a reference to obtain PC1 weights. These findings show that deviations from expected normative microstate architecture are strongly associated with worse treatment outcomes in FEP.

Our findings converge with and extend recent work indicating that microstate architecture capture prognostically relevant variance. De Pieri et al. [22] reported that baseline differences in microstates C, D, and E distinguished subsequent responders from non-responders in multi-episode schizophrenia, while Kikuchi et al.[21] found that changes in microstate D duration correlated with symptom improvement in a small sample. Our study advances this literature by examining minimally treated first-episode patients under standardized treatment conditions—the clinical context where predictive biomarkers would be most valuable—and by employing a normative modeling framework that addresses the analytical challenges inherent in microstate analysis. Importantly, we replicated the pattern of D to C dissociation in schizophrenia compared to healthy controls already described in the literature. Three meta-analyses have been conducted on the topic, with the same directionality of associations (i.e., more presence of C and lower presence of D), where class C increase is driven by occurrence and coverage, while class D decreases are driven by reduced duration. One proposed functional interpretation of these changes is an imbalance between salience-related processing (class C, increased) and processes integrating contextual information/ executive control (class D, reduced), consistent with aberrant salience and impaired context updating in psychosis[12–14].

The PCA-based approach we adopted offers several advantages over analyzing individual microstate parameters. Microstate analysis generates highly correlated features creating substantial risk of overfitting and inflated false positive rates when each is tested individually. By deriving a single composite measure from healthy controls and projecting patients onto this normative space, we reduced dimensionality while preserving interpretability. The resulting PC1 captured a pattern of microstate organization dominated by microstates C and F, with loadings reflecting the balance between salience/default mode network activity (microstate C) and posterior-central configurations (microstate F) versus auditory and attention network activity (microstates A and D). This composite measure may better capture the distributed network dysfunction relevant to treatment response than any single microstate parameter.

Notably, the PC1 normative space was transportable between 2 healthy control datasets obtained in different settings (i.e., Germany vs United States), which was further validated by the fact that PC1 in FEP were predictive of treatment response, regardless of the normative dataset used as reference.

At the composite PC1 level responders were statistically indistinguishable from healthy controls, while non-responders showed significant elevation. This suggests that non-response may be characterized by greater deviation from normative brain organization, while patients who respond to first-line antipsychotics may have relatively preserved microstate dynamics. This interpretation aligns with conceptualizations of treatment-resistant schizophrenia as a neurobiologically distinct subtype. For instance, in the MRI imaging domain, deviation from healthy controls has been associated with lower chance of treatment response, including measures such as brain gyrification[38], striatal volume[39], or functional striatal connectivity [40].

The predictive performance of PC1 (AUC = 0.648 for PC1 alone, 0.723 with covariates) represents a modest but statistically significant improvement over chance, as confirmed by permutation testing. This performance is comparable to fMRI based predictive biomarkers [5, 41], and may improve as normative models developed in larger datasets allow to integrate data from additional principal components. Notably, treatment-naïve status—a potential confounder given that prior antipsychotic exposure could influence both microstate properties and treatment response—did not differ between responders and non-responders and was not a significant predictor in adjusted models, strengthening confidence that our findings reflect intrinsic neurobiological differences rather than medication effects. The lack of association between changes in PC1 between baseline and week 12 and symptom change suggest that normative deviation from microstate organization may be a ‘trait’ more than a state biomarker, or that if changes occur in this measure over time they would be of small or moderate magnitude, as our analyses were powered to detect moderate to large effects in this association. This interpretation is supported by the observation that baseline PC1 predicted symptom improvement even within the longitudinal subsample (ρ = 0.34, p = 0.040). If microstate organization reflects relatively stable individual differences in large-scale network dynamics— perhaps influenced by neurodevelopmental factors or illness-related structural changes—it may serve as a prognostic rather than pharmacodynamic biomarker.

Our findings should be interpreted considering several methodological considerations. First, our normative PCA model was derived from a relatively modest healthy control sample (n=34), and only PC1 met our bootstrap stability criterion for retention. While our robustness testing confirmed that PC1 was stable, larger normative samples would likely yield additional stable PCs by virtue of greater statistical power, and therefore could be added to predictive models. Future studies should establish normative microstate models in larger, multi-site healthy control cohorts, which would enable incorporation of multiple principal components and potentially capture additional dimensions of microstate organization relevant to treatment response. Second, our clinical trial employed only two antipsychotic medications (risperidone and aripiprazole), which could limit generalizability. However, medication type was not a significant predictor in any of our models, and both drugs are first-line agents with distinct receptor binding profiles (risperidone being a dopamine D2/serotonin 5-HT2A antagonist and aripiprazole a partial D2 agonist). The consistency of findings across these pharmacologically distinct agents suggests that microstate-based prediction may reflect general antipsychotic responsiveness rather than medication-specific effects, though this requires confirmation with other antipsychotic agents. Third, our cross-validation was performed within-sample which does not substitute for true external validation. The generalizability of our predictive model to independent cohorts, different clinical settings, and diverse patient populations remains to be established. Future studies should prioritize out-of-sample validation in geographically and demographically distinct cohorts. Fourth, our longitudinal sample was powered to detect only medium-to-large associations between change in PC1 and symptom improvement. The lack of statistical significance in the association between change in PC1 and change in symptoms by week 12 does not exclude the possibility of smaller associations that our study was insufficiently powered to detect. While the pattern of results suggests that microstate organization functions as a trait-like predictive marker rather than a state-dependent pharmacodynamic biomarker, this interpretation requires confirmation in larger longitudinal samples adequately powered to detect small effect sizes.

The clinical implications of our findings, while preliminary, are noteworthy. If replicated, microstate-based prediction could inform personalized treatment decisions in first-episode psychosis, potentially identifying patients who may benefit from alternative treatment strategies or earlier escalation to clozapine. The scalability advantages of EEG—including low cost, ease of implementation, and high patient acceptability—make microstate analysis particularly attractive for community mental health settings where most patients with psychosis receive care. However, substantial work remains before clinical implementation, including external validation, establishment of optimal cutoffs, and demonstration of clinical utility in prospective trials.

In conclusion, this study demonstrates that deviation from normative microstate organization at baseline predicts response to first-line antipsychotics in first-episode psychosis. The finding that responders are comparable with healthy controls while non-responders show elevated PC1 suggests that preserved microstate dynamics may be a ‘trait’ marker of treatment responsiveness. These findings advance our understanding of the neurophysiological correlates of antipsychotic response and highlight the potential of EEG microstate analysis as a scalable approach for personalized medicine in early psychosis, while underscoring the need for replication and external validation before clinical implementation.

## Author contributions

Data acquisition: RC, AKM; Analytic design: MM, MS, WV, JMR; Interpretation of the data: JMR, MM, WV, RC, TL, MB, DO, MS, JMK, AKM; First manuscript: JMR. All authors approve the final manuscript.

## Funding/Grant Support

Supported by the National Institutes of Health (K23MH127300 to JR, and R01MH108654 to AKM). No funding was specifically received for this article.

## Conflicts of interest

Dr. Rubio has been a consultant for Lundbeck, Teva, Janssen, Karuna, and Bristol-Myers Squibb, has received research funding by Alkermes and receives royalties from UpToDate. Dr Kane has been a consultant for or received honoraria from Alkermes, Allergan, Boehringer Ingelheim, Cerevel, Dainippon Sumitomo, H. Lundbeck, HealthRhythms, HLS, Indivior, Intracellular Therapies, Janssen Pharmaceutical, Johnson & Johnson, LB Pharmaceuticals, Merck, Minerva, Neurocrine, Newron, Novartis, Otsuka, Roche, Saladax, Sunovion, and Teva. Dr. Kane has received grant support from Otsuka, Lundbeck, Sunovion and Janssen. Dr. Kane is a shareholder in Vanguard Research Group, North Shore Therapeutics, Health Rhythms, MedinCell, and LB Pharmaceuticals, Inc. The rest of the authors have no conflicts to disclose.

## Notes

### Clinical Trial

NCT02822092

### Author Declarations

All participants provided written informed consent under a protocol approved by the Institutional Review Board (IRB) of the Feinstein Institutes for Medical Research at Northwell Health, and the data were part of NIH funded project R01MH108654.

